# Evaluation of the potential incidence of COVID-19 and effectiveness of contention measures in Spain: a data-driven approach

**DOI:** 10.1101/2020.03.01.20029801

**Authors:** Alberto Aleta, Yamir Moreno

## Abstract

Our society is currently experiencing an unprecedented challenge, managing and containing an outbreak of a new coronavirus disease known as COVID-19. While China - where the outbreak started - seems to have been able to contain the growth of the epidemic, different outbreaks are nowadays being detected in multiple countries. Much is currently unknown about the natural history of the disease, such as a possible asymptomatic spreading or the role of age in both the susceptibility and mortality of the disease. Nonetheless, authorities have to take action and implement contention measures, even if not everything is known. To facilitate this task, we have studied the effect of different containment strategies that can be put into effect. Our work specifically refers to the situation in Spain as of February 28th, 2020, where a few dozens of cases have been detected. We implemented an SEIR-metapopulation model that allows tracing explicitly the spatial spread of the disease through data-driven stochastic simulations. Our results are in line with the most recent recommendations from the World Health Organization, namely, that the best strategy is the early detection and isolation of individuals with symptoms, followed by interventions and public recommendations aimed at reducing the transmissibility of the disease, which although not efficacious for disease eradication, would produce as a second order effect a delay of several days in the raise of the number of infected cases.

## I. INTRODUCTION

The first report by the Chinese authorities of the COVID-19 outbreak appeared in December 31st, 2019. Ever since then, the World Health Organization (WHO) and national public health authorities have been tracing and reporting on the evolution of the outbreak. As initially feared, and despite contention measures adopted in China, with a big city like Wuhan being quarantined for weeks, the disease spread beyond mainland China. As of February 29th, 2020, there are 85,403 cases worldwide, of which 79,394 correspond to China [1]. Additionally, 53 countries, including Spain, have reported at least one case of COVID-19. Two months into the outbreak, much is still unknown about the natural history of the disease and the pathogen. Important from the modeling perspective, for instance, it has been claimed that a large number of cases might have gone undetected by routinely screening passengers, due to the special characteristics of this disease [2]. Admmitedly, several studies predict that only between 10% and 20% of the cases have been detected and reported [3–6].

As with any other novel disease, governments, public health services and the scientific community have been working towards stopping the spreading of COVID-19 as soon as possible and with the lowest possible impact on the population [4, 7–9]. From a scientific point of view, there are two course of actions that can be followed. On the one hand, new vaccines and pharmaceutical interventions need to be developed. This usually requires months of work. Therefore, on the other hand, it is important to study the large-scale spatial spreading of the disease through mathematical and computational modeling, which allows evaluating “in-silicon” what-if-scenarios and potential contention measures to stop or delay the disease. This modeling effort is key, as it can contribute to maximize the effectiveness of any protection measures and gain time to develop new drugs or a vaccine to protect the population. Here, we follow the modeling path and analyze, through a data-driven stochastic SEIR-metapopulation model, the temporal and spatial transmission of the COVID-19 disease in Spain as well as the expected impact of possible and customary contention measures.

Our model allows to implement and quantify the impact of several conventional measures in Spain. These policies are mostly aimed at reducing the mobility of individuals, but we also include other plausible settings like a reduction in the time for case detection and isolation. Our findings agree with previous results in the literature that have reported that a reduction as large as 90% in traffic flow has a limited effect on the spreading of the disease. Important enough and at variance with such studies, the data-driven nature of this study and the available dataset allowed us to disentangle the impact of each transportation mode in several scenarios of mobility reduction in Spain. We found that while shutting down completely any transportation means does not lead to a significant reduction in the incidence of the disease, in some contexts the arrival of the peak of the disease is delayed by several days, which could eventually be advantageous. Altogether, we provide evidences supporting the adoption of a mixed strategy that combines some mobility restrictions with, mainly, the early identification of infected individuals and their isolation. These conclusions agree with the latest recommendations by the WHO [10]. We also highlight that although this study has been made with data from Spain, our findings can also be valid for any other country given the ubiquity of mobility patterns worldwide.

## II. MODEL

Stochastic SEIR-metapopulation models are routinely used to study the temporal and spatial transmission of diseases like the COVID-19. Here, we made use of such class of models and implement a data-driven version that allows to obtain realistic estimates for the spatial incidence of the disease as well as its temporal dynamics. More specifically, in terms of time, we feed the model with the available data as of February 28th, 2020. Spatially, we consider that each province (there are 52 in Spain, see appendix B) is represented by a subpopulation. Furthermore, metapopulation models are composed by two types of dynamics: the disease dynamics governed by the chosen compartmental model, SEIR in our case, and the mobility of the individuals across the subpopulations that make up the whole metapopulation system. The latter ingredient, the mobility, connects the subpopulations and allows the disease to spread from one subpopulation to another. In what follows, we describe these two components of our model.

### A. Mobility dynamics

To model the mobility of individuals we use a data-driven approach. Data-driven modeling, at variance with more theoretically inspired methods, has the advantage that it allows to directly implement and evaluate realistic contention measures, thus producing scalable and actionable what-if-scenarios. To this end, we have obtained the inter-province mobility flows provided by the Ministry of Development of Spain [11]. Therefore, the minimal spatial unit in our system is a province. Using the information from the mobility matrices, that report the origin and destination (OD) of individual movements, at each time-step, we sample the number of individuals on the move from each province and distribute them across the country according to the information contained in these OD matrices. Important enough, this dataset not only includes the total number of individuals going from province to province, but it also distinguishes the main transportation means used by the individuals, see Figure 1. This will allow us to gauge the effect of travel restrictions on different transportation modes.

**FIGURE 1:**
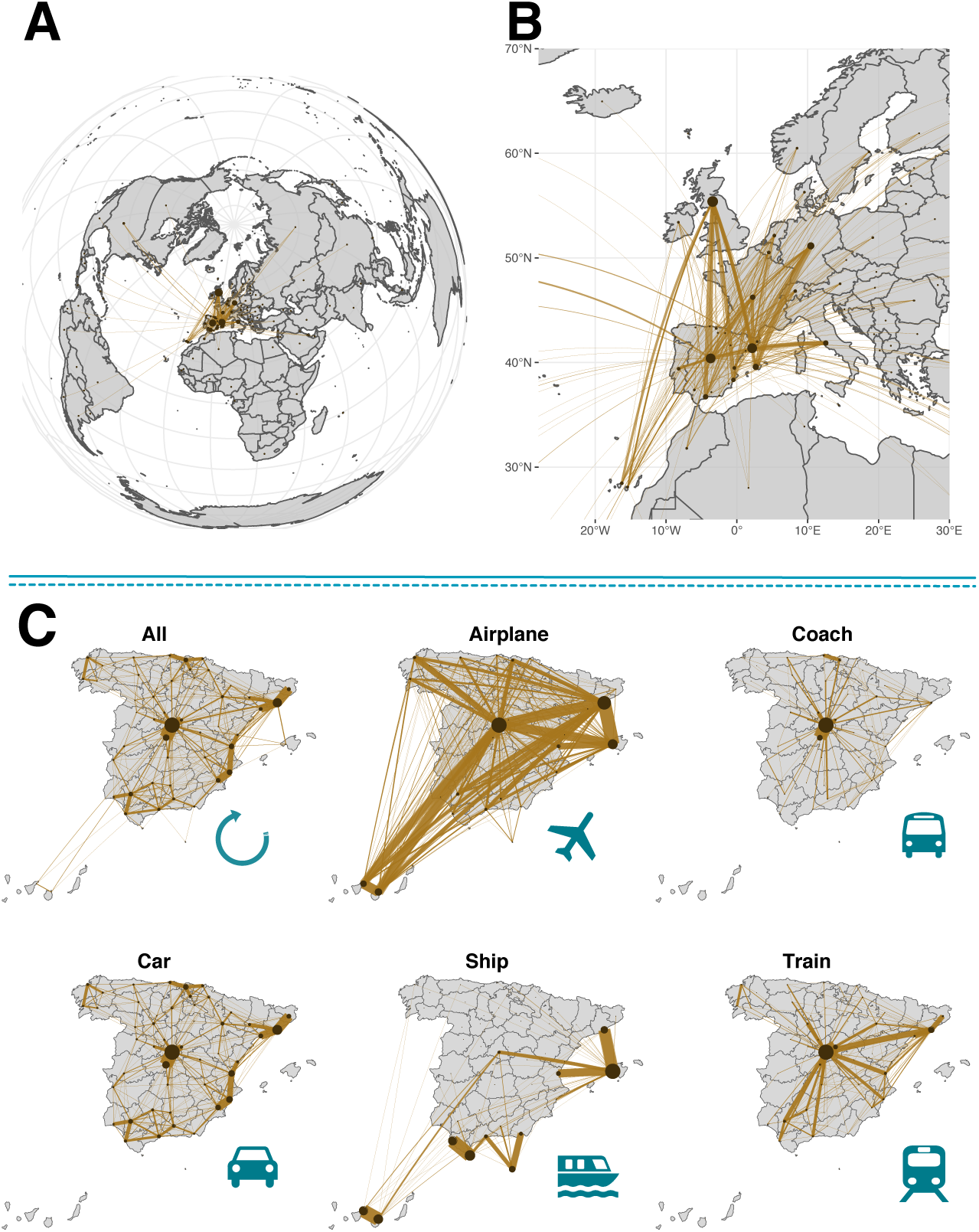
Mobility dynamics for Spain. We use a dataset that includes all possible transportation means, from airplanes to cars. Panels (A) and (B) show the international fluxes to Spain, whereas (C) shows a breakdown of inter-province flows in Spain by transportation mode. The size of the nodes is proportional to the number of individuals leaving the province. Similarly, the width of the links is proportional to the number of individuals using that route. Note that for multimodal travels the associated mode is the one that corresponds to the largest part of the trip, which explains why there are links from the islands to provinces without ports in the matrix corresponding to maritime trips.

Furthermore, given that the epidemic started abroad, it is important to determine in which province the disease is more likely to be seeded first. Given the current global spread and incidence of the epidemic, we thus take into account that the three countries with more cases are China, South Korea and Italy and consider that the most plausible route for an infected individual to reach Spain is by plane. Thus, we collected the number of passengers coming from each country to each Spanish airport in 2019 from the Spanish air navigation manager AENA [12]. Then, we assigned each Spanish airport to its corresponding province and ranked them according to their total number of operations with each country, see figure 2. It is worth noticing that the information provided by AENA is already aggregated by country. Thus, this ranking cannot take into account which airports are mostly connected to locations where the outbreak is currently concentrated − e.g., north of Italy. Nevertheless, the ranking constitutes a valid proxy, and a good starting point, to seed the disease.

**FIGURE 2:**
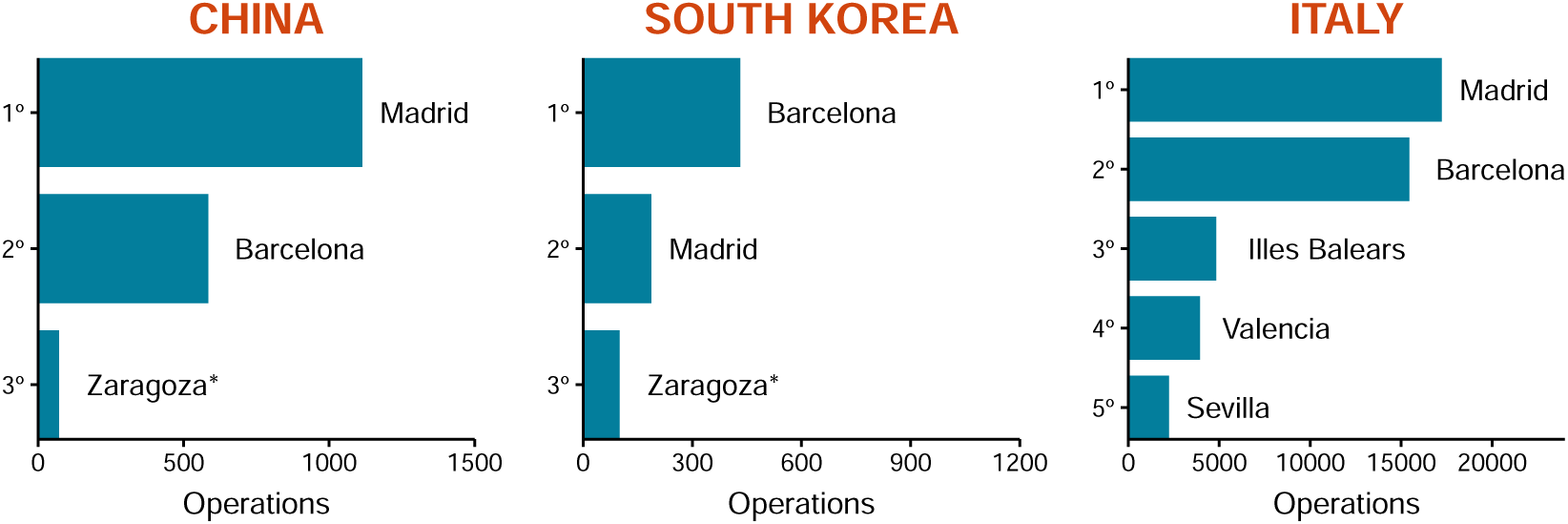
Number of operations (both passengers and cargo) in 2019 from any airport in China, South Korea and Italy to each Spanish airport. Only Madrid and Barcelona have direct passenger connections to China and South Korea, whereas Zaragoza only has freight connections to such locations. The destination provinces are ranked according to the likelihood of receiving an infected individual from each country, assuming the order is proportional to the total number of operations.

### B. Disease dynamics

The dynamics of the disease is governed by an SEIR compartmental model. In this model, individuals are classified according to their health status: susceptible (S) if they are susceptible to catch the disease; exposed (E) if they have been infected but are still asymptomatic and cannot infect other individuals; infected (I) once the incubation period is over and the individuals show symptoms and could infect others, and removed (R) when they are either recovered or deceased. Within each province, the transition between compartments results from the following rules, iterated at each time step, corresponding to 1 day:

**S***→* **E:** Susceptible individuals in province *i* are infected with probability 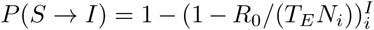, where *R*_0_ is the reproduction number, *T*_*E*_ is the mean incubation time, *N*_*i*_ stands for the number of individuals in region *i* and *I*_*i*_ accounts for the number of infected individuals in such region.

**E***→***I:** Exposed individuals become infected at a rate inversely proportional to the mean latent period, *T*_*E*_.

**I***→***R:** Infected individuals become removed at a rate inversely proportional to the mean infectious period, *T*_*I*_.

In what follows, we parameterize the model according to the latest estimates for the disease parameters [4, 13], namely, *R*_0_ = 2.5, and a generation time *T*_*g*_ = *T*_*E*_ + *T*_*I*_ = 7.5 resulting from considering *T*_*E*_ = 5.2 and *T*_*I*_ = 2.3 (in Appendix D we report that similar results are obtained for other values of *T*_*g*_, see Figures 11, 12 and 13). Additionally, note that we have not explicitly included in the model the possibility of asymptomatic transmission, which should not be mistaken by transmission from undetected or unreported cases. Asymptomatic spreading is still under scrutiny and the statistical evidences are scarce and not significant enough as to be taken by granted. In any case, including this possibility would be trivially possible in this kind of models.

## III. RESULTS AND DISCUSSIONS

### A. Quantifying the spatial and temporal evolution of the disease incidence

One of the main characteristics of the COVID19 disease is its long latency times. Currently, the average incubation period has been reported to be 5.2 [13]. Thus, before proceeding with the evaluating the impact of the disease, we first compare the metapopulation model employed here with a classical SIR-metapopulation framework. To do so, we use the random-walk effective distance

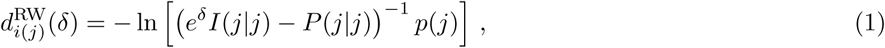

where *P* (*j* |*j*) is the normalized flow matrix without row and column *j, p*(*j*) is the *j*-column of *P* with element *j* removed and *δ* is a dimensionless parameter that depends on the infection, recovery and mobility rates [14]. This quantity, defined for SIR metapopulation models, gives us the expected time that it would take for the disease to reach each subpopulation of the system, also known as the hitting time. In Figure 3, we compare the hitting time obtained from stochastic simulations of the SEIR metapopulation model with the theoretical distances derived for the simplified SIR model. We can see that the correlation is nearly perfect, implying that the spreading itself is quite similar in both models. However, we find that the hitting times in the SEIR implementation are at least two times larger than the theoretical ones for the SIR scenario (on its turn, stochastic simulations of the SIR model agree very well with the theoretical expectations for the model, see appendix C). Thus, the addition of the latent state produces a substantial delay on the spreading of the epidemic. This is in line with the fact that the epidemic is thought to have started in mid-November or early December, however, a noticeable number of cases was only reported by early January.

**FIGURE 3:**
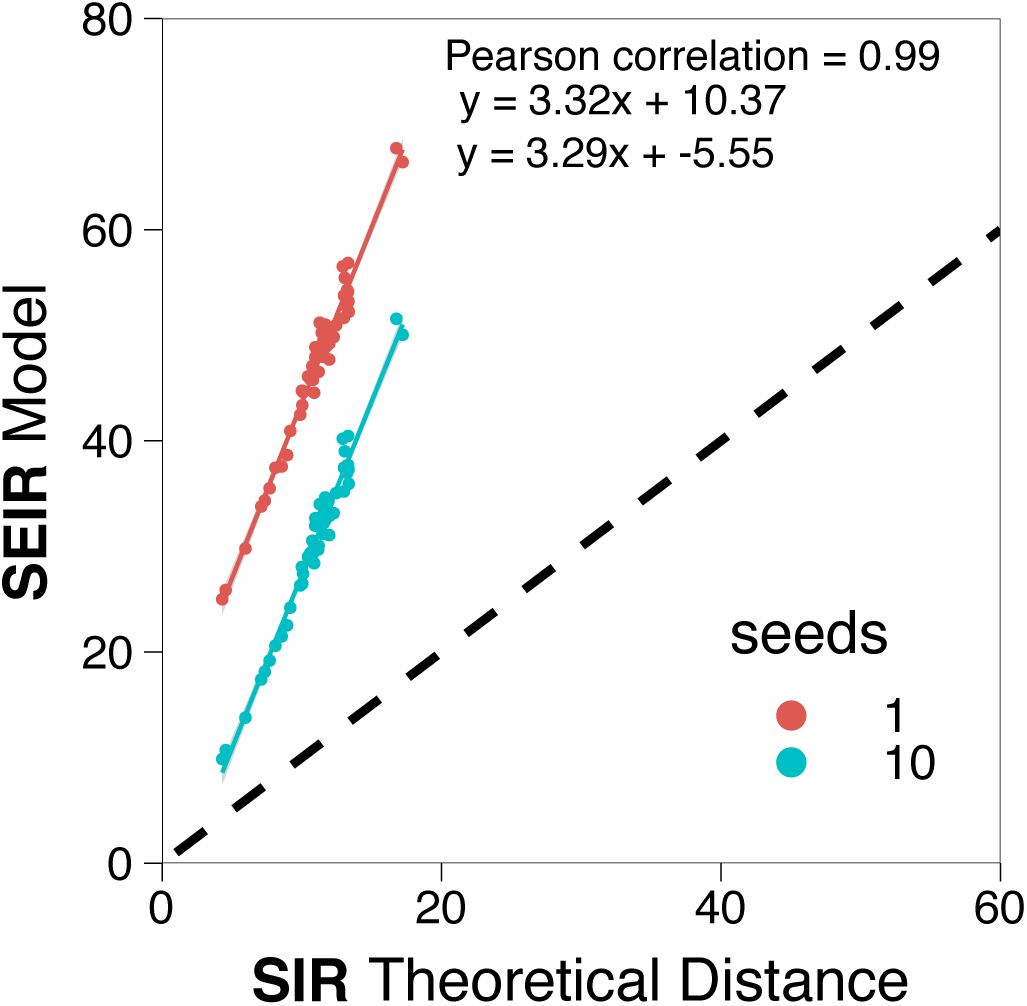
Comparison between the hitting time obtained after 10^3^ simulations of the SEIR model in our metapopulation scheme, with 1 or 10 seeds initially placed in the province of Barcelona, and the theoretical distance in an SIR metapopulation model.

Figure 4 shows the expected hitting time for each province when the disease starts from 5 different locations, as well as one case with seeds in multiple places, as obtained from the SEIR metapopulation model. As before, the hitting times might seem long, but this is due to the long latent periods of the disease, in agreement with the evolution of reported cases in mainland China. We also note that Spanish major cities are expected to be affected by the outbreak in no more than 40 days − but often within 20 days − from the initial time in all the situations considered. Indeed, to mimic the situation in the country as of February 28th, 2020, and to make projections about the evolution of the disease into the future, we considered the scenario in which the model is initialized with infected individuals in the capital of each region where there are cases of COVID19: Madrid, Tenerife, Barcelona, Balearic Islands, Zaragoza, Seville and Valencia [15]. The results show that the spreading is much faster (note that the outer circle in the 5 individual provinces represents 60 days while in this case it represents only 40 days) in such a situation.

**FIGURE 4:**
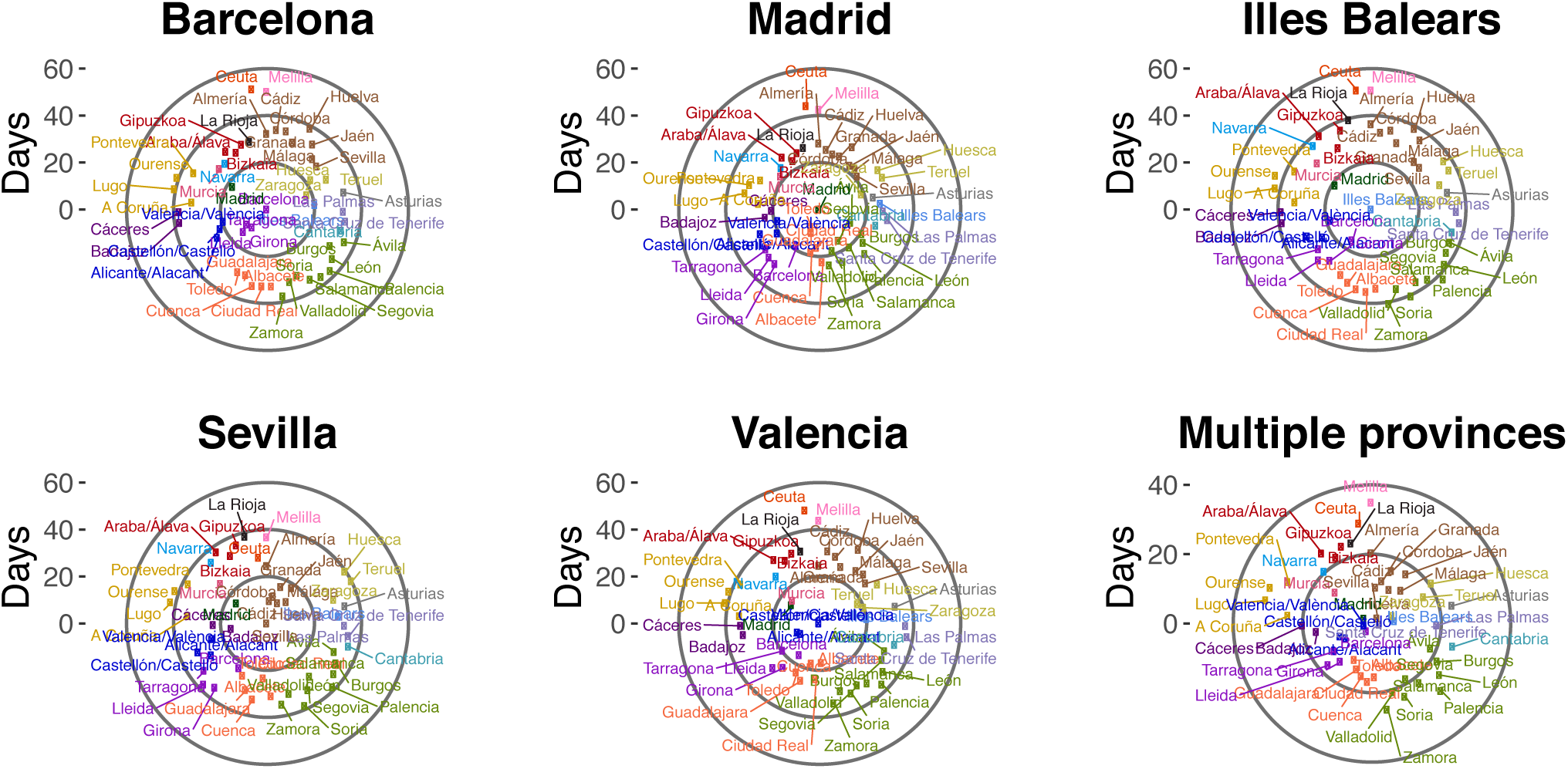
Time needed for an infected individual to arrive to each province if the disease starts with 10 infected individuals in Barcelona, Madrid, Illes Balears, Sevilla or Valencia. In the multiple provinces case, we introduced 5 in Madrid, 6 in Tenerife, 4 in Barcelona, 1 in Illes Balears, 1 in Zaragoza, 8 in Sevilla and 10 in Valencia. As indicated in each plot, we have simulated scenarios in which this number of infected individuals is seeded in five different major cities in Spain as well as for the case in which there are multiple places seeded at the same time.

Complementarily to Figure 4, we present in Figure 5 further results on the temporal and spatial evolution of the disease dynamics. Here we have computed, through stochastic simulations of the model, the cumulative number of infected individuals within each region assuming that the disease propagates from Barcelona by 10 initially infected individuals. The results align with the theoretical predictions, and highlight the close relationship that exists between the two biggest cities of Spain (Barcelona and Madrid), even though they are relatively far geographically (around 620 kilometers through the shortest path by car). Again, note that the relative slow speed of the epidemic agrees with what has been observed in China. Finally, it is worth remarking that all of these quantities have been obtained assuming that no control measures are taken, which is not the current case as (detected) infected individuals are being isolated and monitored by the authorities. Additionally, we also stress that it is difficult to assess whether this corresponds to a worst-case projection or not, given the many unknowns that can’t be taken into account, such as inflow of infected subjects from abroad, the fraction of infectious individuals that have gone undetected, etc. However, as we show next, this data-driven modeling approach do allow to evaluate the effect of customary contention measures.

**FIGURE 5:**
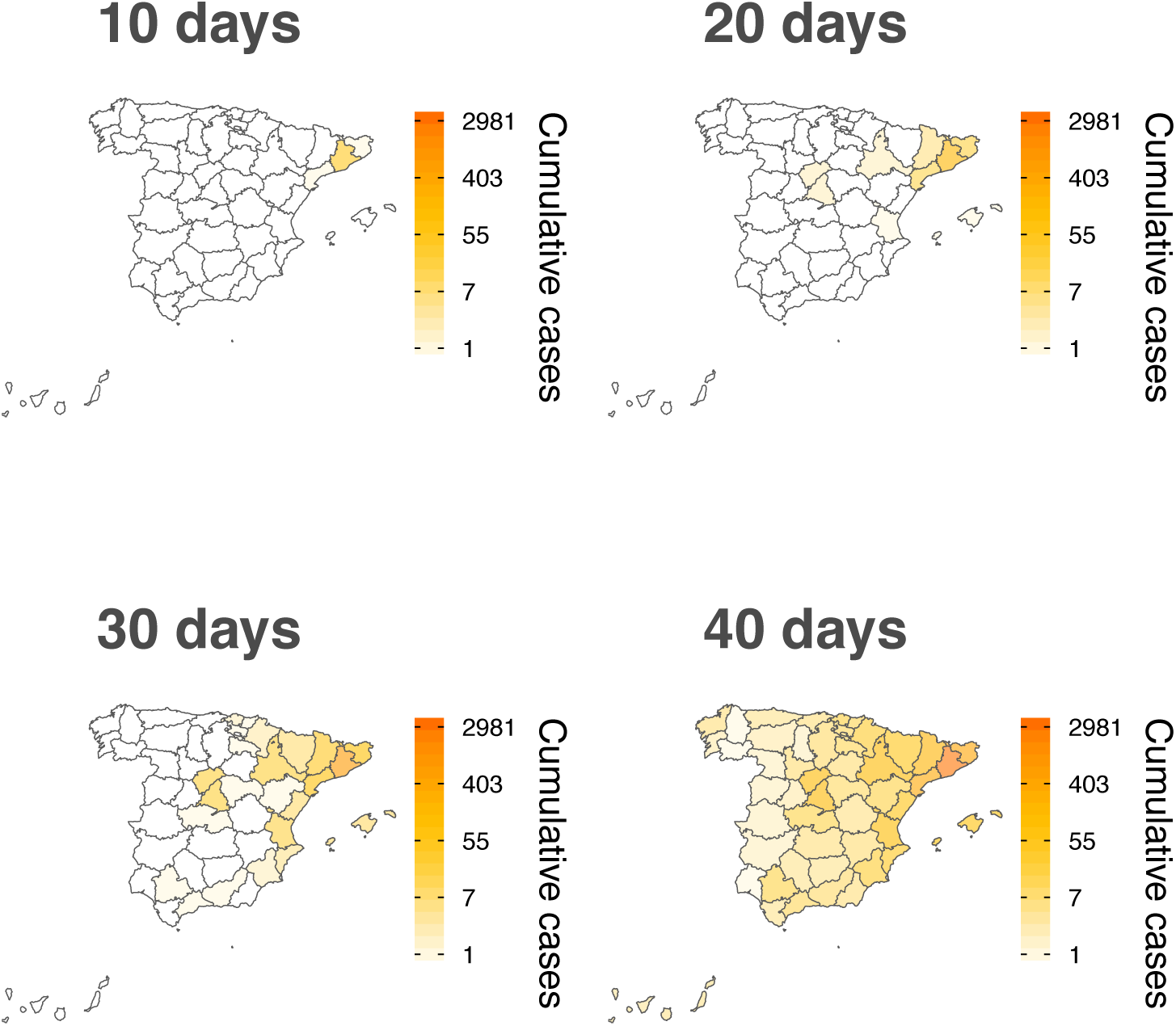
Estimated cumulative number of infected individuals within each region when the disease starts with 10 infected individuals in the province of Barcelona. The reported values are the median over 10^3^ simulations.

## IV. CONTAINMENT OF THE EPIDEMIC

Our data-driven model is particularly useful to get insights into mobility-mediated transmission dynamics and to evaluate possible countermeasures. Next, we explore diverse containment measures that could be implemented aiming at stopping the large-scale spreading of the disease. First, we analyze the effect of imposing mobility restrictions by limiting traffic flow in the country. We consider six different scenarios that correspond to each transportation mode being shutdown plus another one in which a total reduction of 90% of the overall traffic is imposed. These measures are extreme and unless the situation gets really critical, would not be put into practice as they bear an economic cost that would be insurmountable. Nonetheless, as we show below, however drastic they appear to be, these measures are useless when it comes to completely stop the disease from propagating. Indeed, a significant reduction in the estimated incidence is only obtained when other actions are feasible.

Admittedly, in Figure 6A it is observed that the previous measures have no effect on the final size of the epidemic. On the other hand, if we look at the time for the peak of the epidemic to arrive, Figure 6B, we see some minor effects. In particular, although shutting down most modes of transportation have practically no effect, if all private cars were removed (i.e., they remain confined in their corresponding province), the peak of the epidemic would be delayed by about 7 days. The most effective of the above scenarios of mobility restriction corresponds to an unrealistic 90% reduction of the overall traffic, when the peak would be delayed over 20 days. This is in agreement with previous studies that have shown that the only sizable effect of travel restrictions is to delay the peak of the epidemic. For instance, it has been claimed that the travel restrictions in Wuhan only delayed the peak of the epidemic by 3 days [4].

**FIGURE 6:**
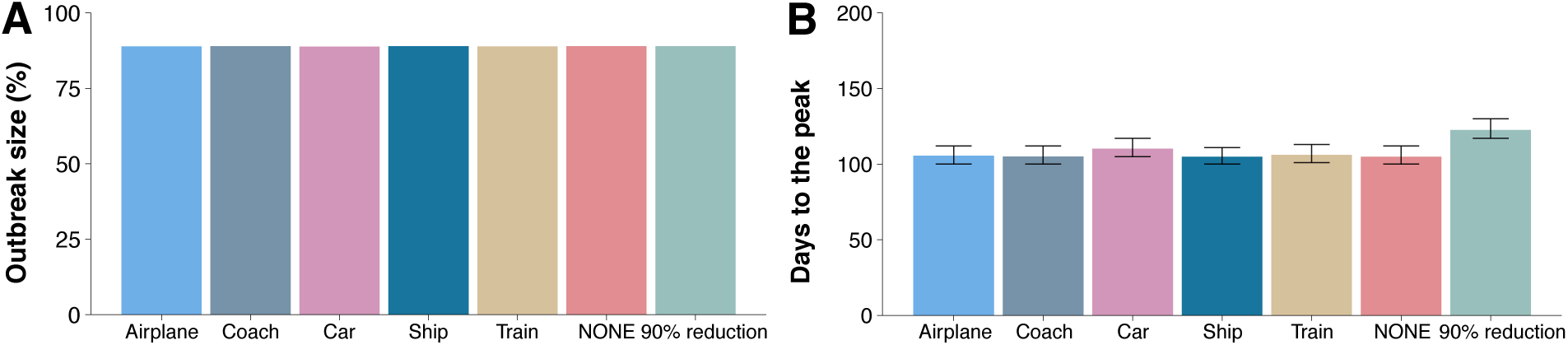
Evaluating the impact of mobility reduction. Panel (A) shows the fraction of individuals who where affected by the disease by the end of the epidemic, whereas panel (B) displays the time from the arrival of the first infected individual to the country until the peak of the epidemic, i.e., the day with the maximum number of infected individuals. In all cases the epidemic starts with 10 infected individuals places in Barcelona.

Another possibility, instead of limiting the mobility of the overall population, is to be extremely vigilant so as to make it possible to isolate all the cases that start to appear quickly enough. To check the impact of this policy, we have simulated a scenario in which the average number of days that an individual is able to go unnoticed and infect others is reduced from 2.3 to 2, 1.5 and 1 days. In Figure 7A, we observe that this strategy is much more effective than traffic restrictions. In particular, if we were able to reduce the time since symptoms’ onset to isolation below 1.5 days, the epidemic is greatly reduced. As a matter of fact, it has been recently reported [13] that this average number of days went down in China from 4.4 days at the beginning of the outbreak to 2.6 days, which is one of the reasons invoked to explain why the epidemic has started to decline in mainland China. In our case, these numbers would be compatible with generation times of 10 or 12.5 (see figure 12).

**FIGURE 7:**
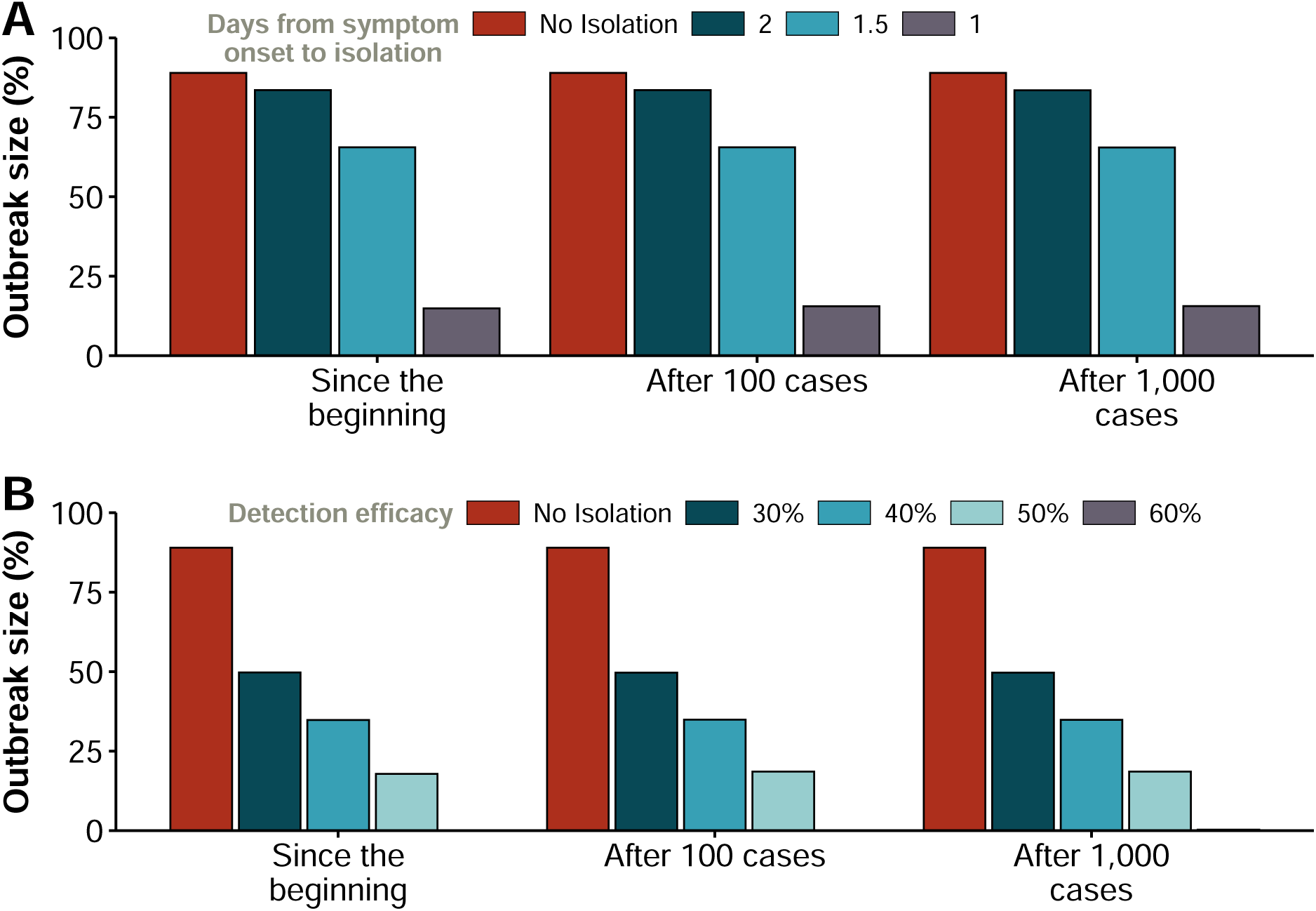
Size of the epidemic if (A) individuals are hospitalized or isolated after a given number of days from the onset of disease symptoms; and (B) a certain fraction of individuals is hospitalized or isolated after they experience the first symptoms. Each set of measures indicates when they were applied: since the beginning of the epidemic, and after 100 or 1,000 cases are detected in the whole country. Note that individuals who escape isolation remain infectious until they recover, and thus they could be thought of as asymptomatic spreaders.

It is also reasonable to assume that this strategy is not easy to implement in full, either because some individuals could purportedly try to avoid isolation or due to the fact that many infected subjects have mild symptoms similar to a common flu and neither go to the doctor nor report their state. Therefore, we have simulated a slightly different scenario in which individuals are isolated the same day of their symptoms’ onset with a certain efficacy. That is, only a given percentage of the new cases is isolated, while the others are able to roam freely. This framework would also be compatible with having asymptomatic individuals who are able to spread the disease, something that is currently under debate and not yet statistically supported. The equivalence with such hypothetical natural history of the disease in our model is such because we do not apply the prescribed percentage to the total number of infected individuals, but only to those who have just developed symptoms, thus, those that escape will remain infectious as if they were asymptomatic until they recover. In Figure 7B we show the effect that different percentages of new isolated cases would have on the size of the outbreak. Being able to hospitalize all individuals, on average, in less than 1 day enables to effectively stop the disease. Yet, the results also show that even if all infectious are not isolated, as long as more than 60% of the infected individuals are, the effect would be similar and the disease would be effectively eradicated. Lastly, we analyzed the consequences of self-protection measures such as wearing masks, washing more frequently ones hands or avoiding crowded places. To mimic these contexts, we simply reduced the effectivity of the transmission by a certain fraction, and study the final size of the epidemic, see Figure 8A. The results show that a large reduction of at least 60% is needed to contain the disease. Interestingly, if we look at the time to the peak of the epidemic, represented in Figure 8B, we observe that decreasing the transmission not only reduces the size of the outbreak but also delays the peak. Hence, even if this strategy might not be sufficient to completely stop the propagation of the disease in all cases, it could certainly help for preparedness and other clinical responses by delaying the spreading. The exception is when the reduction is very large (in the figure, beyond 60%) as in these cases the peak might occur earlier because the disease dies out.

**FIGURE 8:**
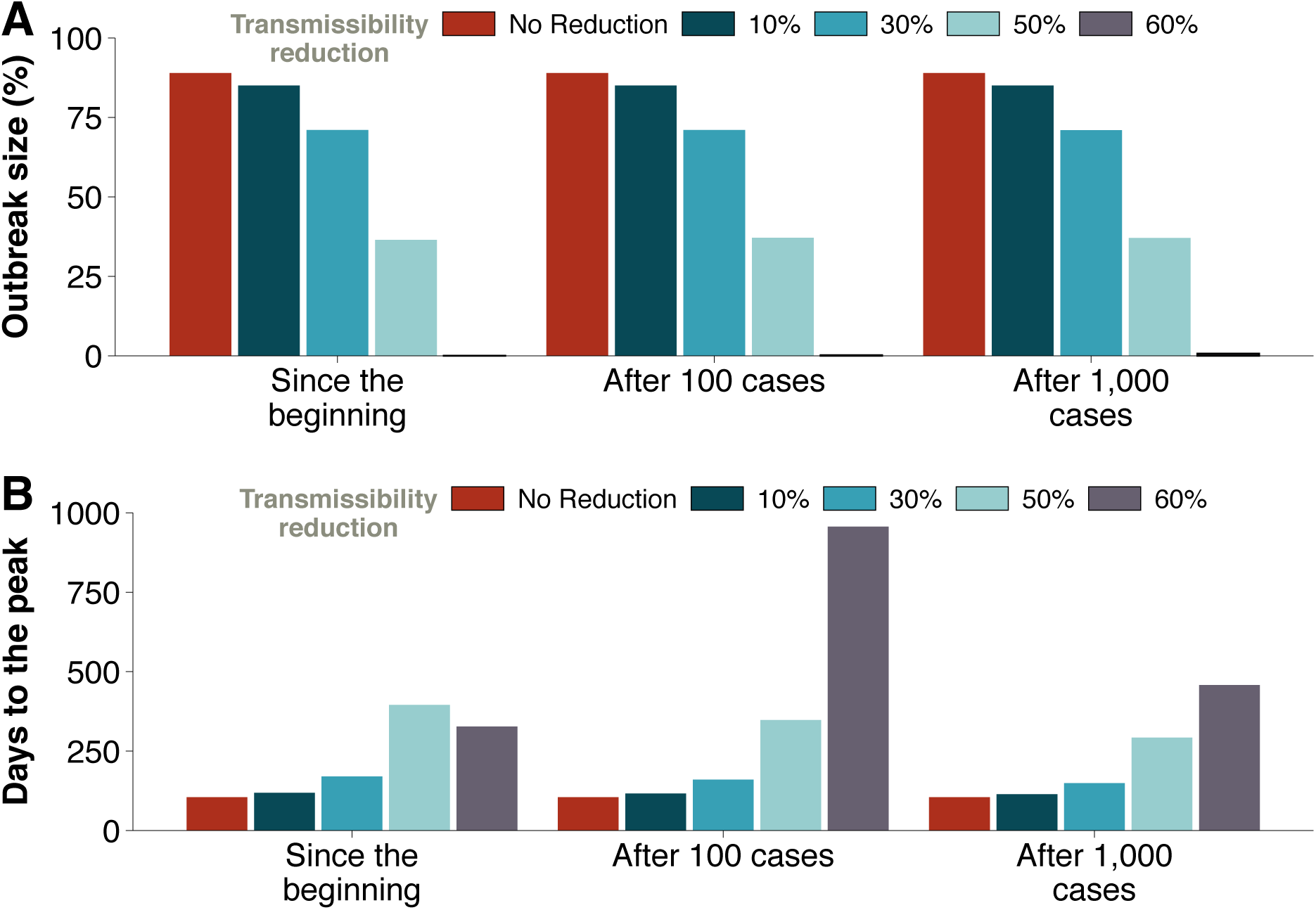
Effect of a reduction in transmission on the (A) outbreak size and (B) time for the disease to peak. The simulated reductions are as indicated in the legends and have been implemented either from the onset of the outbreak or when a certain number of cases in the whole country are detected. Once the transmissibility is reduced by 60% or more, the epidemic fades out. Note that reducing the transmissibility always delays the spreading, except in situations where the disease dies out, for which the peak occurs earlier.

**FIG. 9:**
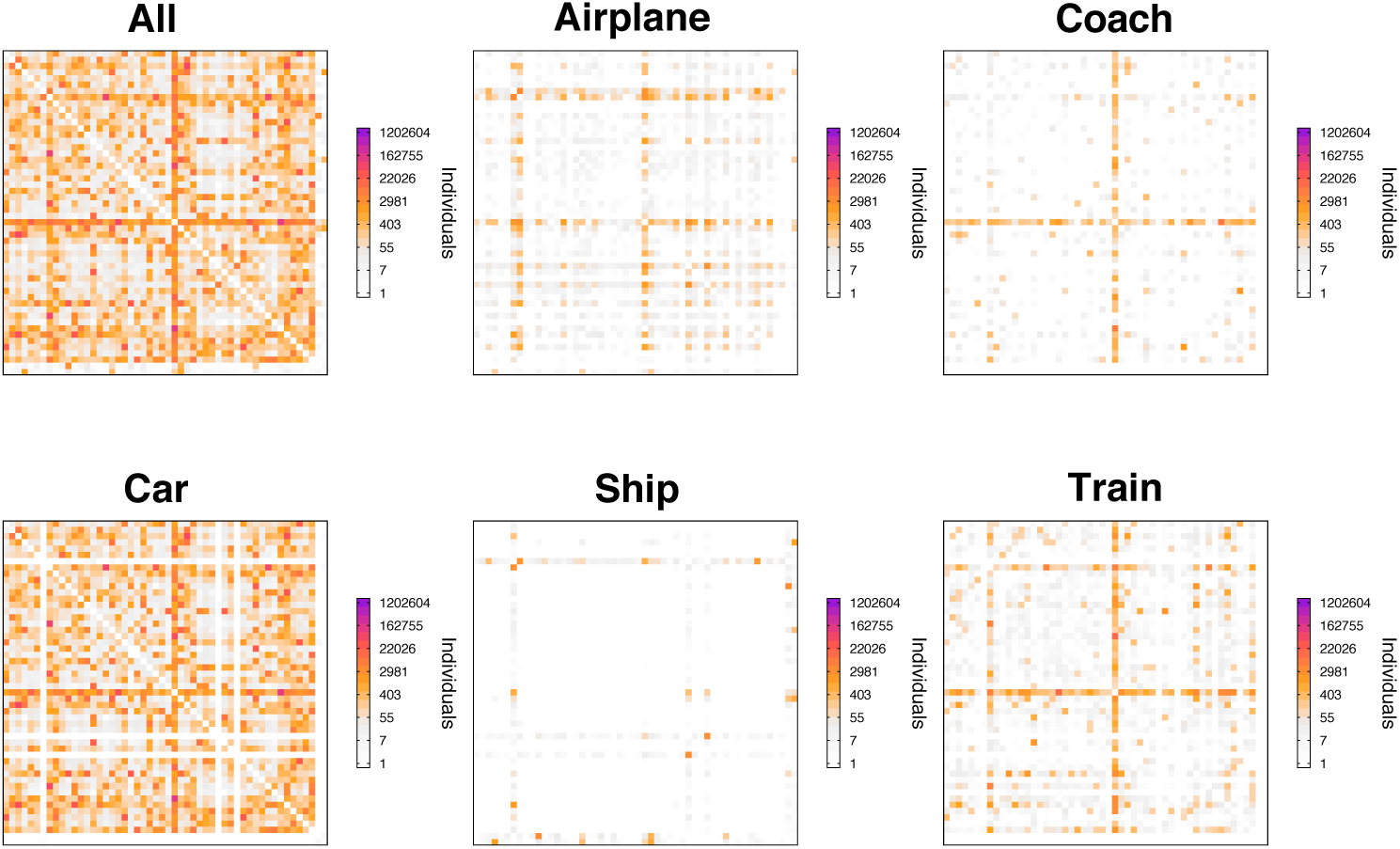
Average ow in matrices in the last two weeks of October 2017.

In concluding, it is apparent from all the results obtained for the different scenarios that we have considered that the most cost-effective strategy would be the isolation or quarantine of detected infectious cases, as long as the efficacy of such measure is over 60%. Important for the current debate about the natural history of the disease, this policy would also work if there is a fraction of asymptomatic but infectious individuals in the population. Our results also show that from a practical point of view, a combination of all the analyzed contexts can have second order benefits. As already stressed, containment measures should not only be directed towards a full cutdown of the number of infected cases. Their efficacy is also given by other factors, such as delaying, even if only by a few days, the spreading of the disease. Such delays are always good for preparedness and to have more time for clinical research that can lead to new pharmacological treatments or vaccines. For instance, even if traffic restrictions are not effective on their own, they facilitate the control of the population and thus it would be easier to detect infected individuals and treat them. Similarly, self-protection measures and other social-distancing practices delay the spreading of the disease, freeing resources that would allow for a better management of the epidemic, in turn leading to an increase of the efficacy of individual isolation. Closing public places would, in practice, reduce the transmission, which again will lower the total number of infections and thus make them more manageable for the public health system. This also highlights the importance of having a coordinated response system, since simply adopting central measures like imposing mobility restrictions and closing public places is not effective in the middle-to-long term.

Our model has several limitations and some of them could actually be overcome in the near future. Perhaps the most important one has to do with the inability of current large-scale epidemiological models to fully account for behavioral changes in the population when a disease is evolving. As it is the case for the spreading of COVID-19, the information − and more often than desired, misinformation− travels faster than the disease. This produces undesired effects such as a collapse in the emergency rooms at hospitals, a proliferation of information sources that do not provide sensible advices in all cases, anticipated economic loses and, in general, uncoordinated responses. Therefore, it is a pressing challenge to develop more realistic ways to incorporate in models like the one employed here all these risk-averse responses and reactions. Another limitation of the current study includes the relatively low spatial resolution, which is essentially determined by the data availability. The results however indicate that the level of granularity used here is enough to capture mobility patterns and the effects of possible interventions. Finally, we have not considered the temporal and spatial variability of disease parameters, nor other potentially important characteristics of the host population such as the existence of super spreaders or the age structure, which seems to play a relevant role for this disease, at least in what concerns the case fatality rate. We plan to investigate on all these issues in the near future.

## Data Availability

Data is freely available from the authors or the sources reported in the manuscript.

## Acknowledgments

YM acknowledges partial support from the Government of Aragon, Spain through grant E36-17R (FENOL), and by MINECO and FEDER funds (FIS2017-87519-P). AA and YM acknowledge support from Intesa Sanpaolo Innovation Center. The funders had no role in study design, data collection, and analysis, decision to publish, or preparation of the manuscript.

## APPENDIX A MOVEMENT DATA

The inter-province flow matrices include the number of individuals that move from province to province in Spain for several days of October 2017. The data also includes the main mode of transport used by those individuals, as well as other characteristics such as the period of the day when the travel started. In our case, we have collected the matrices from the last two weeks of October and averaged them. For a deeper discussion on the characteristics of these matrices we refer the reader to the original source (in spanish) [11].

## APPENDIX B GEOGRAPHIC AND DEMOGRAPHIC DATA

The upper level of administrative division in Spain is denominated *Comunidad Autónoma*. There are 15 of such “autonomous communities” in mainland Spain, one in the islands of the mediterranean sea (Balearic Islands) and one for the islands in the Atlantic Ocean (Canary Islands). Besides, there are two autonomous cities (*ciudades autónomas*) in the north of Africa, Ceuta and Melilla. The next administrative division is the province (*provincia*). There are 47 provinces in mainland Spain, 1 in the Balearic Islands and 2 in the Canary Islands, plus the two autonomous cities making up the 52 subpopulations considered in our model. The number of inhabitants of each province varies a lot, from over 5 millions in Madrid and Barcelona to less than 100,000 individuals in Ceuta and Melilla. We collected the number of inhabitants of each province from the data provided by the Spanish Statistical Office [16].

There are 45 airports in Spain (including the heliports in Algeciras and Ceuta), although the majority of them only have connections to other Spanish airports or European airports. As with the provinces, the traffic in these airports varies widely, from over 50 million passengers in Madrid and Barcelona to less than 2,000 in Albacete and Huesca. The information regarding Spanish airports is provided by the Spanish air navigation manager (AENA) [12].

## APPENDIX C EFFECTIVE DISTANCES FOR EPIDEMIC SPREADING

To ensure that this quantity is able to predict correctly the spreading in our metapopulation, we first implement an SIR model and study the effect of using a different number of seeds, figure 10. Regardless on the number of seeds, the correlation between the obtained values and the theoretical ones is really large. However, the actual value depends on the number of initial seeds. Thus, without any modification this measure can help us determine the arrival order, i.e., to which provinces the disease should arrive first, but not the precise time. Nonetheless, depending on the number of seeds, a small correction has to be applied.

**FIGURE 10:**
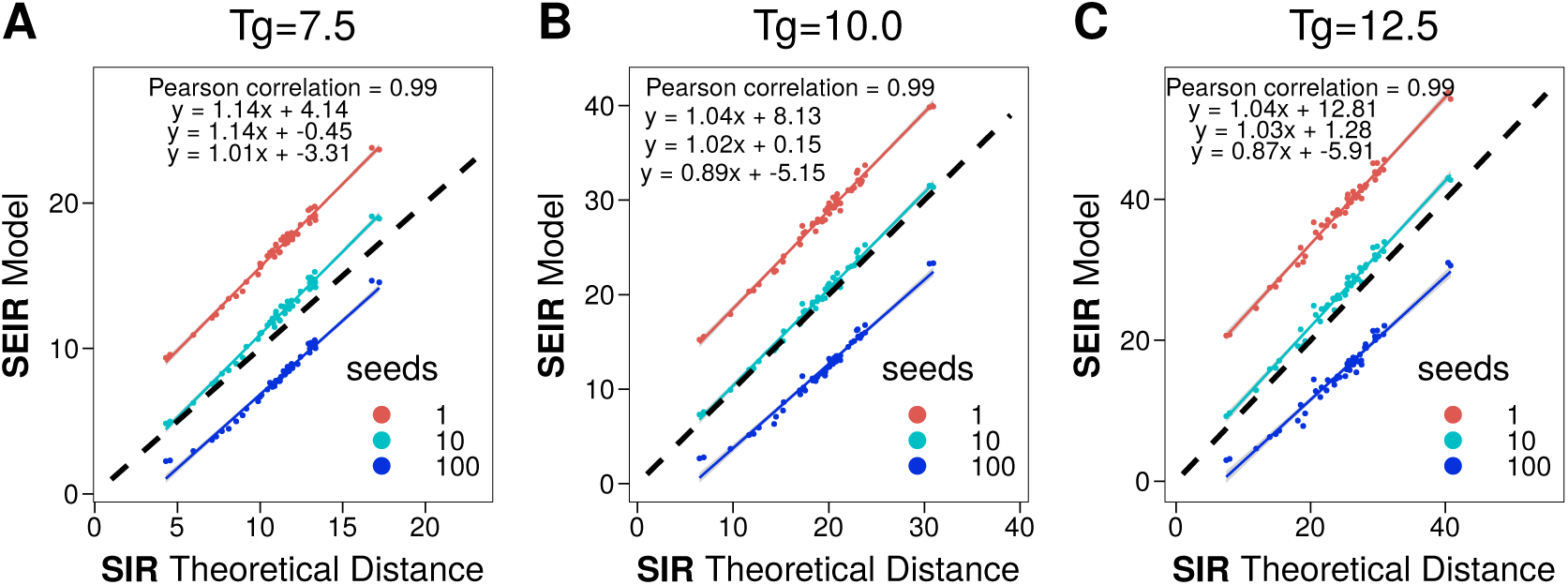
Relationship between the hitting time in a simulation of SIR dynamics in our metapopulation and the theoretical one for different number of seeds and three values of the generation time *T*_*g*_.

## APPENDIX D ROBUSTNESS OF THE RESULTS

As it is not clear yet what are the final disease parameters that affect the predictions about the spreading of the coronavirus, we here provide quantitative evidences that the results reported in the main text still hold for other values of the generation time *T*_*g*_. A similar exercise could be done for *R*_0_, but we believe that the consensus around the value used here (2.5) is higher than for other parameters like the serial interval. Figures 11, 12 and 13 show results obtained for *T*_*g*_ = 10 and *T*_*g*_ = 12.5. As it can be seen, the previous conclusions hold, and in most of the cases, also quantitatively in terms of relative variation with respect to the scenario in which no measures are implemented.

**FIGURE 11:**
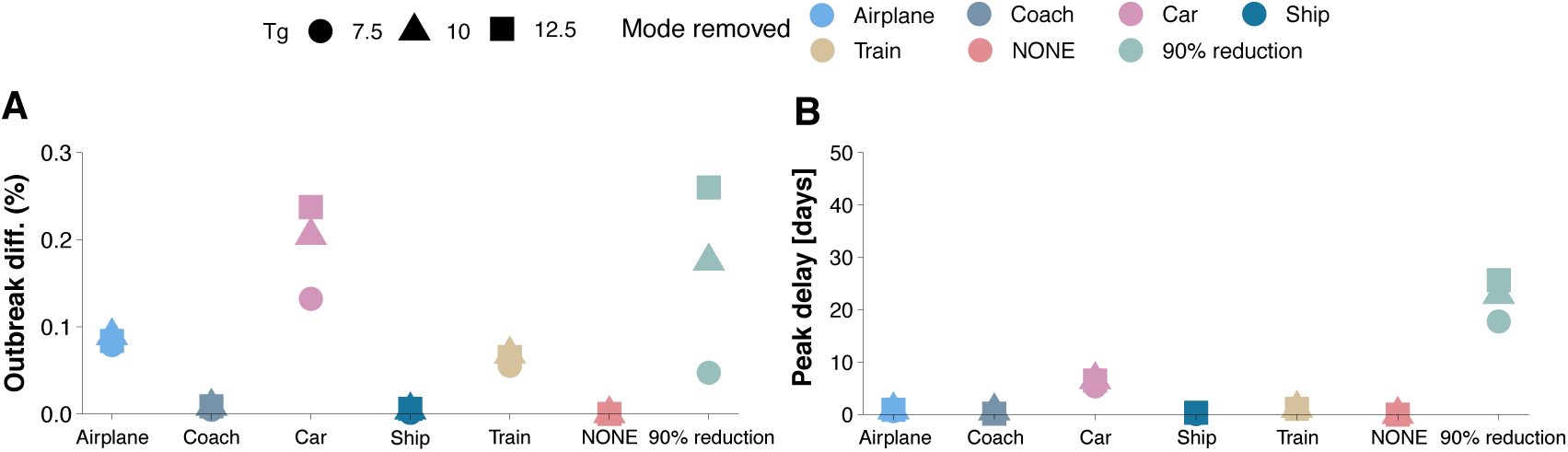
Sensitivity analysis with respect to the value of *T*_*g*_. The figure shows the results displayed in Figure 6 but including *T*_*g*_ = 10 and *T*_*g*_ = 12.5. *R*_0_ = 2.5.

**FIGURE 12:**
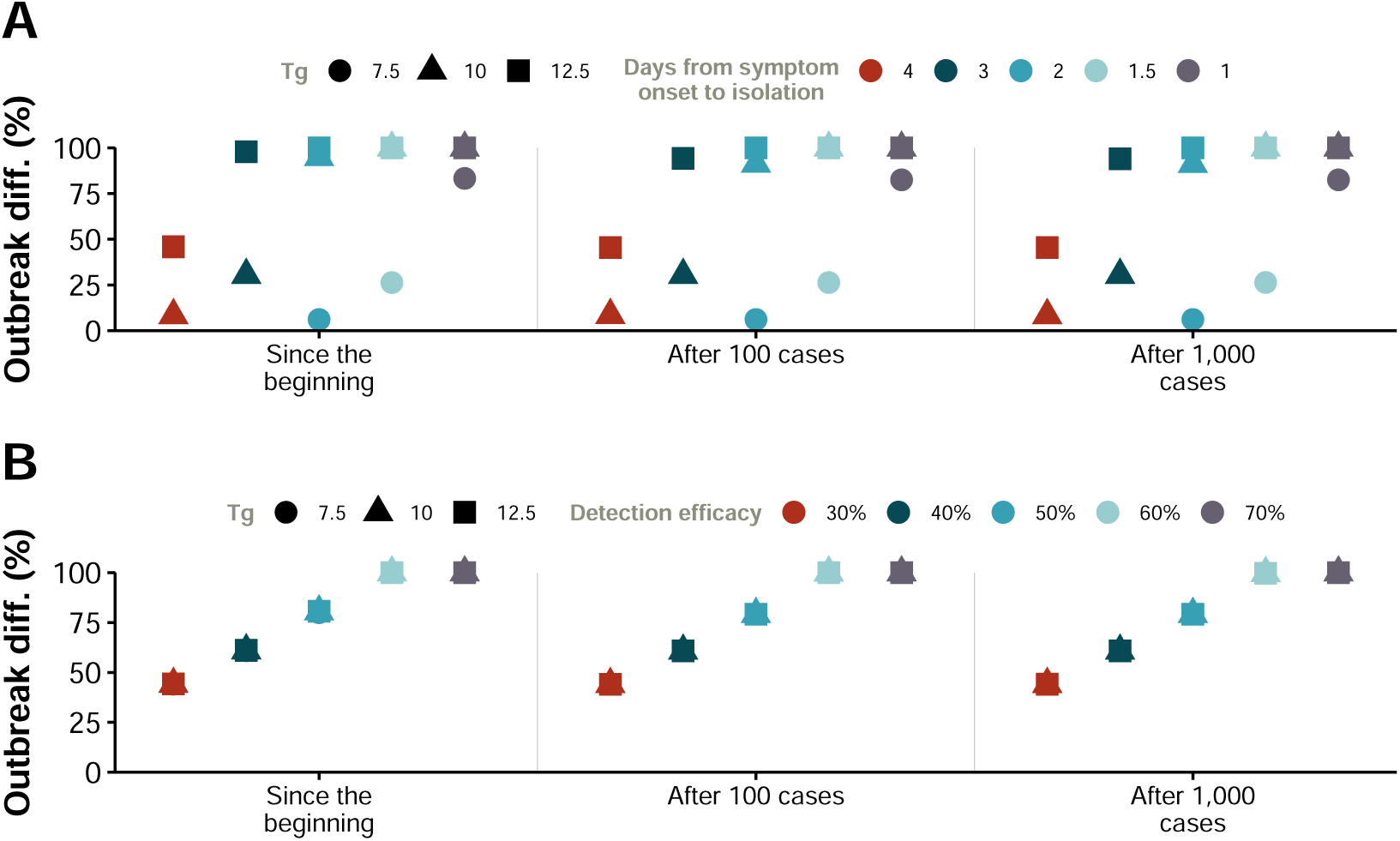
Sensitivity analysis with respect to the value of *T*_*g*_. The figure shows the results displayed in Figure 7 but including *T*_*g*_ = 10 and *T*_*g*_ = 12.5. *R*_0_ = 2.5.

**FIGURE 13:**
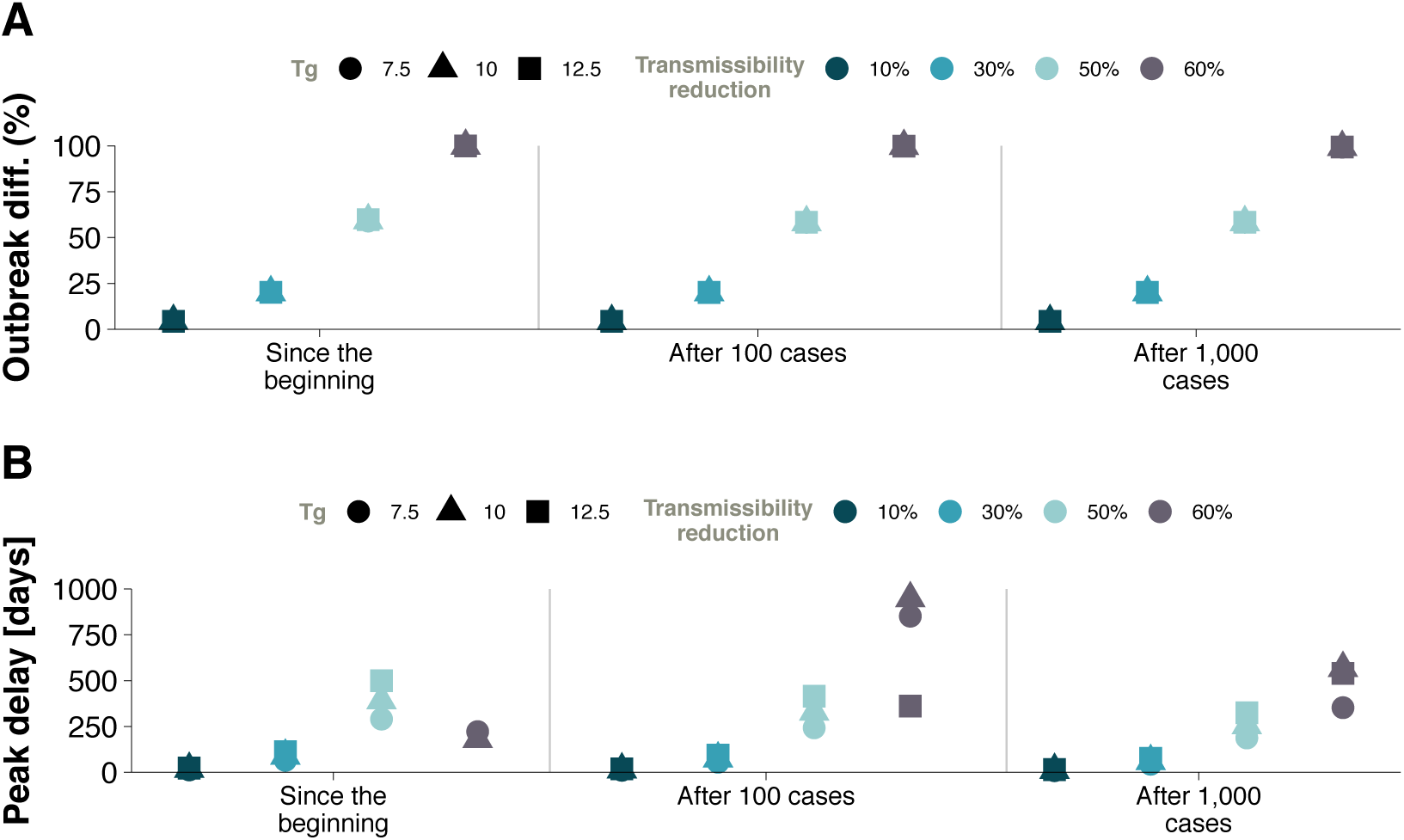
Sensitivity analysis with respect to the value of *T*_*g*_. The figure shows the results displayed in Figure 8 but including *T*_*g*_ = 10 and *T*_*g*_ = 12.5. *R*_0_ = 2.5.

## References

[1] Tech. Rep., World Health Organization (2020), URL https://www.who.int/docs/default-source/coronaviruse/situation-reports/20200229-sitrep-40-covid-19.pdf?sfvrsn=7203e653_2.

[2] K. Gostic, A. C. R. Gomez, R. O. Mummah, A. J. Kucharski, and J. O. Lloyd-Smith, eLife (2020).

[3] Z. Du, L. Wang, S. Cauchemez, X. Xu, X. Wang, B. J. Cowling, and L. A. Meyers, medRxiv p. 2020.01.28.20019299 (2020).

[4] M. Chinazzi, J. T. Davis, M. Ajelli, C. Gioannini, M. Litvinova, S. Merler, A. Pastore y. Piontti, K. Mu, L. Rossi, K. Sun, et al., Science p. eaba9757 (2020), ISSN 0036-8075.

[5] Z. Cao, Q. Zhang, X. Lu, D. Pfeiffer, L. Wang, H. Song, T. Pei, Z. Jia, and D. D. Zeng, medRxiv p. 2020.02.07.20021071 (2020).

[6] Y. Zhou and J. Dong, medRxiv p. 2020.02.10.20021774 (2020).

[7] X. Zhu, A. Zhang, S. Xu, P. Jia, X. Tan, J. Tian, T. Wei, Z. Quan, and J. Yu, medRxiv p. 2020.02.09.20021360 (2020).

[8] X. Li, X. Zhao, and Y. Sun, medRxiv p. 2020.02.09.20021477 (2020).

[9] H. Xiong and H. Yan, medRxiv p. 2020.02.10.20021519 (2020).

[10] Tech. Rep., World Health Organization (2020), URL https://www.who.int/docs/default-source/coronaviruse/who-china-joint-mission-on-covid-19-final-report.pdf.

[11] Tech. Rep., Ministry of Development of Spain (2019), URL https://observatoriotransporte.mitma.gob.es/estudio-experimental.

[12] Spanish air navigation manager, AENA (acceseed 2020), URL http://www.aena.es.

[13] J. Zhang, M. Litvinova, W. Wang, Y. Wang, X. Deng, X. Chen, M. Li, W. Zheng, L. Yi, X. Chen, et al., medRxiv p. 2020.02.21.20026328 (2020).

[14] F. Iannelli, A. Koher, D. Brockmann, P. Höovel, and I. M. Sokolov, Phys. Rev. E 95, 012313 (2017), URL https://link.aps.org/doi/10.1103/PhysRevE.95.012313.

[15] Regions with cases of COVID19 as of 29/02 in Spain (acceseed 2020), URL https://elpais.com/sociedad/2020-02-28/los-contagios-activos-por-coronavirus-se-elevan-a-30-en-espana.html.

[16] Spanish Statistical Office (in spanish) (accessed February 2020), https://www.ine.es/.

